# Rising Prevalence of Cardiovascular-Kidney-Metabolic (CKM) Syndrome in China, 2010-2019: National Cross-Sectional Surveys

**DOI:** 10.1101/2025.04.21.25323810

**Authors:** Daijun He, Xiao Zhang, Chenglong Li, Chao Yang, Mengting Yu, Jinwei Wang, Jiakang Huo, Bixia Gao, Ming-hui Zhao, Mei Zhang, Limin Wang, Luxia Zhang, Erdan Dong

## Abstract

**Background:** Cardio-kidney-metabolic (CKM) syndrome is defined as a systemic disorder characterized by pathophysiological interactions among metabolic risk factors, chronic kidney disease, and the cardiovascular system, leading to multiorgan dysfunction and a high rate of adverse cardiovascular outcomes. However, the prevalence of CKM syndrome among adults in China is unknown.

**Objectives:** The aim of this study was to assess the prevalence of CKM syndrome and its stages in mainland China.

**Methods:** This cross-sectional study included two nationally representative data from 41 088 participants aged 18 and older from the China National Survey of Chronic Kidney Disease (CNSCKD) conducted from September 2009 to September 2010 and 171 462 adults from the sixth China Chronic Disease and Risk Factor Surveillance (CCDRFS) conducted from August 2018 to June 2019. CKM syndrome stages were identified in accordance with the 2023 American Heart Association Presidential Advisory on CKM Health. Prevalence estimates were weighted to represent the Chinese adult population accounting for the complex sampling design. The prevalence of advanced CKM syndrome was compared among subgroups using log-binomial regression with age and sex adjusted. Weighted multivariable logistic regression models for advanced CKM syndrome were analyzed with relevant covariates.

**Results:** The weighted prevalence of CKM syndrome and its advanced stages among adults in mainland China were 77.1% (95% confidence interval, 76.1% to 78.0%) and 6.3% (5.9% to 6.7%) in 2010, and 83.7% (83.2% to 84.2%) and 10.1% (9.9% to 10.4%) in 2019. Among all CKM syndrome stages, stage 2 had the highest prevalence, accounting for 50% of the population in both 2010 and 2019. The prevalence of CKM syndrome have increased across all age groups, especially the 18-39 age group. The increase was greater in men than women across all age groups. Factors independently associated with advanced CKM syndrome were age, sex, area of residence, education level, smoking status, alcohol consumption, diet, and physical activity.

**Conclusions:** The prevalence of CKM syndrome in China is high and has increased significantly over the past decade, emphasizing the urgent need to develop comprehensive and equitable prevention and management strategies.

## Introduction

Globally, the prevalence of obesity has increased steadily, with 43% of the world’s population now classified as overweight or obese.(1) It is widely recognized as a major risk factor for several chronic diseases, including diabetes, cardiovascular disease (CVD), and chronic kidney disease (CKD).(2,3) Concurrently, the incidence of diabetes, CVD, CKD, and related comorbidities has also escalated, contributing to substantial rises in morbidity, mortality, and healthcare expenditures.(4–7) These chronic conditions not only overlap within the population but also share common pathophysiological pathways, including dysglycemia, dyslipidemia, and hypertension.(8) Consequently, impairment in one system may promote and amplify dysfunction in others, leading to further morbidity and mortality.(8) For instance, in a nationwide sample, although diabetes and CKD were each associated with high 10-year cardiovascular mortality rates (6.7% and 9.9%, respectively), the combination of diabetes and CKD was linked to a synergistically higher 10-year cardiovascular mortality rate (19.6%).(9) As our understanding of these diseases deepens, the shared risk factors and interactions among the cardiovascular system, kidneys, and metabolism have become increasingly recognized. Without timely and comprehensive management, these interactions can lead to a self-reinforcing cycle that accelerates disease progression.

To facilitate interdisciplinary care of these diseases, the American Heart Association (AHA) issued a presidential advisory introducing the novel concept of cardio-kidney-metabolic (CKM) syndrome in 2023.(10) CKM syndrome is defined as a systemic disorder characterized by pathophysiological interactions among metabolic risk factors, CKD, and the cardiovascular system, leading to multiorgan dysfunction and a high rate of adverse cardiovascular outcomes.(11) China, as a rapidly developing low- and middle-income country, is experiencing a steady rise in the prevalence of non-communicable diseases and associated metabolic risk factors, driven by profound socioeconomic changes.(12) In China, the estimated prevalence of individual CKM conditions were 50.7% for overweight or obese, 23.4% for CVD, 10.8% for CKD, and 11.2% for diabetes.(13–15) Despite the sustained growth in the burden of CKM conditions in China, the prevalence of CKM syndrome remains largely unknown. Understanding its prevalence and possible trends is essential for supporting evidence-based public health policies and developing a multidisciplinary approach to management and prevention.

In this study, two nationally representative data from the China National Survey of Chronic Kidney Disease (CNSCKD) conducted from September 2009 to September 2010 and the sixth China Chronic Disease and Risk Factor Surveillance (CCDRFS) conducted from August 2018 to June 2019 were used to examine the prevalence of CKM syndrome and its potential trends among Chinese adults.

## Methods

### Study population

The CNSCKD recruited a representative sample of Chinese adults in mainland China using a multistage, stratified sampling method and was conducted from September 2009 to September 2010. Details of the study design have been previously described.(14) 50 550 people were invited to participate and 47 204 completed the survey and examination, with a response rate of 93%. In the current analysis, participants with missing information on variables for evaluating CKM syndrome (n=6116) were excluded. Overall, 41 088 participants from the CNSCKD were included (supplementary fig 1).

The sixth CCDRFS was a large, cross-sectional, nationally representative survey that covered all 31 provincial-level administrative divisions in mainland China and was conducted from August 2018 to June 2019. Details of the study design have been previously described.(16) A total of 184 876 eligible adults agreed to participate in the survey, with a response rate of 94.9%. In the current analysis, individuals with missing information on variables for evaluating CKM syndrome (n=13 414) were excluded. Overall, 171 462 participants from the sixth CCDRFS were included (supplementary fig 1).

The study protocol was approved by the ethics review committee of the National Center for Chronic and Noncommunicable Diseases Control and Prevention, and Peking University First Hospital. All participants provided written informed consent before data collection.

### Data Collection

In the CNSCKD, all participants completed a questionnaire documenting their sociodemographic status, personal health history, and lifestyle with assistance from trained personnel. Anthropometric measurements were obtained and body mass index (BMI) was calculated as weight in kilograms divided by the square of height in meters. Blood and urine samples were analyzed at central laboratories in each province, all of which successfully completed a standardization and certification program.

In the sixth CCDRFS, each participant completed a comprehensive questionnaire by trained personnel on sociodemographic information, behavioral and dietary habits, physical activity, medical history, and a physical examination including height, weight, waist circumference (WC), and blood pressure. BMI was calculated as weight in kilograms divided by height in meters squared. Blood samples after an overnight fast of at least 10 hours and morning urine samples were collected from all participants. Assay for fasting blood glucose and 2-hour oral glucose tolerance test (OGTT) was completed within 24 hours of the testing in 298 local laboratories that had passed a technical appraisal. All other assays were completed in 3 central laboratories that had completed a standardization and certification program. Detailed descriptions of data collection in both surveys are provided in the Supplementary Method 1.

### Assessment criteria

CKM syndrome stages were identified among participants in both the CNSCKD and the sixth CCDRFS in accordance with the 2023 AHA Presidential Advisory on CKM Health(10).

CKM Stage 0 included participants with normal BMI (<23 kg/m^2^), normal WC (<80 and <90 cm for women and men, respectively) who did not meet criteria for the other stages.

CKM Stage 1 identified individuals with an elevated BMI (≥23 kg/m^2^), an elevated WC (≥80 and ≥90 cm for women and men, respectively), or prediabetes (defined as a HbA1c of 5.7% to <6.5% or a fasting blood glucose of 100 mg/dL to <126 mg/dL).

CKM Stage 2 was characterized by the presence of metabolic risk factors or moderate-to-high-risk CKD. Metabolic risk factors included elevated fasting serum triglycerides (TG) (≥135 mg/dL), hypertension, diabetes, or metabolic syndrome (MetS, ≥3 of the following: elevated WC, low high density lipoprotein cholesterol (HDL-C) level [<40 mg/dL or <50 mg/dL for men or women, respectively], TG ≥150 mg/dL, elevated blood pressure [systolic blood pressure (SBP) ≥130, diastolic blood pressure (DBP) ≥80 mmHg, and/or use of blood pressure-lowering medications], or fasting blood glucose ≥100 mg/dL). CKD stages were identified based on estimated glomerular filtration rate (eGFR) and urinary albumin-to-creatinine ratio (uACR).(17) eGFR was calculated with 2021 race-free Chronic Kidney Disease Epidemiology Collaboration creatinine equation and uACR was measured with immunoturbidimetric tests in both surveys.(18)

CKM Stage 3 was identified based of the presence of very-high-risk KDIGO CKD stages or a high-predicted 10-year CVD risk. 10-year cardiovascular risk was estimated with the AHA Predicting Risk of CVD EVENTs (PREVENT) equations.(19) High risk was defined as ≥20% 10-year CVD risk. The PREVENT equations were developed and validated for adults 30-79 years of age. As such, risk was not estimated for adults <30 years. However, to minimize underestimation of CKD Stage 3, adults ≥80 years were not excluded from 10-year CVD risk. Instead, adults ≥80 years were assigned an age of 79 years when determining 10-year CVD risk to allow for conservative estimates. Further, PREVENT was developed for variables with the following ranges: total cholesterol (TC) 130-320 mg/dL, HDL-C 20-100 mg/dL, SBP 90-200 mmHg, and eGFR 14-140 mL/min/1.73m². To approximate PREVENT risk strata, values for these variables above or below these bounds were set to the upper or lower bounds of allowable values respectively (for example, TC of 330 mg/dL was set as 320 mg/dL). Cardiac biomarkers and cardiovascular imaging were not available to identify subclinical CVD.

CKM Stage 4 was identified based on self-reported established CVD (myocardial infarction and stroke in the CNSCKD; myocardial infarction, angina, atrial fibrillation, and stroke in the sixth CCDRFS).

Advanced CKM syndrome were defined as stages 3 or 4 because these identify individuals with or at high risk of CVD.

### Statistical analysis

The characteristics of the study population were summarized as categorical variables and presented as proportions. Relevant characteristics are described and stratified according to CKM syndrome stages. The prevalence of CKM syndrome stages in the overall population and different strata were calculated using sample weights that incorporated multistage sampling weight, the nonresponse weight, and the population weight. These weights were used to adjust for different selection probabilities, different response proportions, and deviations in the sample compared with the standard population, particularly for age and sex.(14,20) Data from the China Population Sampling Census in 2009 were used as the standard population in the CNSCKD, and data from the sixth national census in 2010 were used as the standard population in the sixth CCDRFS. The 95% confidence intervals (CIs) of the prevalence estimates were calculated using Taylor series linearization with finite population correction implemented in the proc surveyfreq procedure in SAS, version 9.4 (SAS Institute Inc). The prevalence of advanced CKM syndrome was compared among subgroups using log-binomial regression with age and sex adjusted. Since CKM syndrome stage 2 is primarily defined by the presence of metabolic risk factors, we assessed potential changes in the distribution of these factors within this stage. Weighted multivariable logistic regression models for advanced CKM syndrome were analyzed with relevant covariates. The prevalence of hypertension, diabetes, dyslipidemia, hyperuricemia, and CKD as well as the corresponding rates of awareness, treatment, and control among the treated patients, were also estimated.

Statistical analyses were conducted using SAS 9.4 (SAS Institute, Cary, NC) and R language 4.3.2 (R Foundation, Vienna, Austria), with a two-tailed alpha of 0.05 considered statistically significant.

## Results

### Study Population

Table 1 shows the general characteristics of participants from the CNSCKD and the sixth CCDRFS. Among 41 088 participants from the CNSCKD, 17.2% were 60 years or older, 49.3% were female, 78.5% were from rural areas, and 95.2% were of Han ethnicity. Among 171 462 participants from the sixth CCDRFS, 17.3% were 60 years or older, 49.6% were female, 48.2% were from rural areas, and 91.1% were of Han ethnicity. Adults of advanced CKM syndrome stages were more likely to be older, men, from rural areas, less educated, current smoker, and habitual drinker. Low levels of fruit and vegetable intake, red meat intake, and physical activity were more prevalent among adults of advanced CKM syndrome stages (supplementary table 1).

**Table 1.** Baseline characteristics of participants from the China National Survey of Chronic Kidney Disease (CNSCKD) and the sixth China Chronic Disease and Risk Factor Surveillance (CCDRFS) according to cardiovascular-kidney-metabolic syndrome stages.

| Characteristic | CNSCKD, 2009-2010, No. (%)<sup>a</sup> |  |  |  |  |  | sixth CCDRFS, 2018-2019, No. (%)<sup>a</sup> |  |  |  |  |  |
| --- | --- | --- | --- | --- | --- | --- | --- | --- | --- | --- | --- | --- |
|  | Total<br>(n=41088) | Stage 0<br>(n = 9250) | Stage 1<br>(n = 7900) | Stage 2<br>(n = 20015) | Stage 3<br>(n = 2912) | Stage 4<br>(n = 1011) | Total<br>(n=171462) | Stage 0<br>(n = 16548) | Stage 1<br>(n = 31289) | Stage 2<br>(n = 92013) | Stage 3<br>(n = 16478) | Stage 4<br>(n = 15134) |
| Age group, years |  |  |  |  |  |  |  |  |  |  |  |  |
| 18-39 | 9975 (42.1) | 4337 (61.7) | 2290 (51.6) | 3310 (33.8) | 12 (1.1) | 26 (13.1) | 25467 (44.8) | 6477 (73.2) | 7728 (55.6) | 10995 (38.9) | 35 (1.0) | 232 (8.4) |
| 40-59 | 20225 (40.7) | 3983 (32.0) | 4551 (40.6) | 11218 (48.2) | 183 (6.7) | 290 (38.6) | 76453 (37.9) | 6761 (22.1) | 16034 (36.5) | 48186 (46.5) | 1347 (11.6) | 4125 (35.8) |
| 60-69 | 6738 (10.6) | 716 (4.7) | 831 (5.8) | 4377 (14.0) | 496 (20.6) | 318 (24.0) | 47011 (9.8) | 2626 (3.4) | 5975 (5.7) | 27070 (11.2) | 4636 (17.6) | 6704 (28.0) |
| ≥70 | 4150 (6.6) | 214 (1.6) | 228 (2.0) | 1110 (4.0) | 2221 (71.6) | 377 (24.3) | 22531 (7.5) | 684 (1.3) | 1552 (2.1) | 5762 (3.4) | 10460 (69.7) | 4073 (27.7) |
| Sex |  |  |  |  |  |  |  |  |  |  |  |  |
| Women | 23865 (49.3) | 5885 (48.1) | 4878 (53.1) | 11401 (49.6) | 1207 (36.4) | 494 (46.9) | 95303 (49.6) | 9771 (59.3) | 18784 (56.8) | 53134 (44.8) | 5728 (37.0) | 7886 (46.7) |
| Men | 17223 (50.7) | 3365 (51.9) | 3022 (46.9) | 8614 (50.4) | 1705 (63.6) | 517 (53.1) | 76159 (50.4) | 6777 (40.7) | 12506 (43.2) | 38878 (55.2) | 10750 (63.0) | 7248 (53.3) |
| Township |  |  |  |  |  |  |  |  |  |  |  |  |
| Urban | 21812 (21.5) | 4940 (24.8) | 4124 (17.0) | 10494 (21.5) | 1616 (27.0) | 638 (18.0) | 69997 (51.8) | 6444 (55.3) | 12227 (51.3) | 37831 (51.9) | 6760 (44.9) | 6735 (49.2) |
| Rural | 19276 (78.5) | 4310 (75.2) | 3776 (83.0) | 9521 (78.5) | 1296 (73.0) | 373 (82.0) | 101465 (48.2) | 10104 (44.7) | 19063 (48.7) | 54181 (48.1) | 9718 (55.1) | 8399 (50.8) |
| Geographical region |  |  |  |  |  |  |  |  |  |  |  |  |
| South | 20037 (59.0) | 5170 (65.8) | 3406 (47.9) | 9630 (59.3) | 1443 (73.1) | 388 (58.0) | 108862 (72.2) | 11487 (77.9) | 20742 (73.3) | 57740 (71.0) | 11300 (74.80) | 7593 (58.2) |
| North | 21051 (41.0) | 4080 (34.2) | 4494 (52.1) | 10385 (40.7) | 1469 (26.9) | 623 (42.0) | 62600 (27.8) | 5061 (22.1) | 10548 (26.7) | 34272 (29.0) | 5178 (25.2) | 7541 (41.8) |
| Ethnicity |  |  |  |  |  |  |  |  |  |  |  |  |
| Han | 36849 (95.2) | 7774 (94.2) | 7072 (94.8) | 18321 (95.6) | 2737 (97.2) | 945 (95.2) | 150686 (91.1) | 13788 (90.7) | 26768 (90.9) | 81274 (91.3) | 15100 (92.9) | 13756 (89.8) |
| Other<sup>b</sup> | 4239 (4.8) | 1476 (5.8) | 828 (5.2) | 1694 (4.4) | 175 (2.8) | 66 (4.8) | 20776 (8.9) | 2760 (9.3) | 4522 (9.1) | 10738 (8.7) | 1378 (7.1) | 1378 (10.2) |
| Education |  |  |  |  |  |  |  |  |  |  |  |  |
| High school or above | 17611 (31.4) | 4900 (41.5) | 3775 (32.6) | 8069 (28.8) | 584 (10.5) | 283 (12.7) | 34064 (34.9) | 4747 (49.6) | 7133 (38.5) | 17987 (32.5) | 1874 (12.3) | 2323 (18.3) |
| Secondary school or lower | 23477 (68.6) | 4350 (58.5) | 4125 (67.4) | 11946 (71.2) | 2328 (89.5) | 728 (87.3) | 137398 (65.1) | 11801 (50.4) | 24157 (61.5) | 74025 (67.5) | 14604 (87.7) | 12811 (81.7) |
| Smoking status |  |  |  |  |  |  |  |  |  |  |  |  |
| Current | 9847 (28.4) | 1869 (29.0) | 1702 (25.2) | 4999 (28.3) | 1017 (41.1) | 260 (24.4) | 41650 (26.3) | 4161 (22.4) | 6917 (21.6) | 20530 (28.4) | 6656 (38.6) | 3386 (25.7) |
| Former/Never | 31241 (71.6) | 7381 (71.0) | 6198 (74.8) | 15016 (71.7) | 1895 (58.9) | 751 (75.6) | 129812 (73.7) | 12387 (77.6) | 24373 (78.4) | 71482 (71.6) | 9822 (61.4) | 11748 (74.3) |
| Drinking status |  |  |  |  |  |  |  |  |  |  |  |  |
| Habitual drinker | 3221 (6.8) | 469 (3.5) | 559 (5.7) | 1715 (8.0) | 387 (13.4) | 91 (8.4) | 17306 (8.0) | 1203 (4.2) | 2701 (6.0) | 9152 (8.8) | 2847 (16.7) | 1403 (12.4) |
| Non-habitual drinker | 37867 (93.2) | 8781 (96.5) | 7341 (94.3) | 18300 (92.0) | 2525 (86.6) | 920 (91.6) | 154136 (92.0) | 15343 (95.8) | 28587 (94.0) | 82851 (91.2) | 13627 (83.3) | 13728 (87.6) |

<sup>a</sup> All percentages are weighted.
<sup>b</sup> Other includes all ethnic minorities in mainland China.

### Prevalence of CKM syndrome

In the CNSCKD, the weighted prevalence of CKM syndrome by stage were 22.9% (95% CI, 22.0% to 23.9%) for stage 0, 21.4% (20.5% to 22.3%) for stage 1, 49.4% (48.3% to 50.4%) for stage 2, 4.9% (4.6% to 5.3%) for stage 3, 1.3% (1.1% to 1.6%) for stage 4. In the sixth CCDRFS, the weighted prevalence by stage were 16.3% (15.8% to 16.8%) for stage 0, 22.8% (22.3% to 23.3%) for stage 1, 50.8% (50.1% to 51.4%) for stage 2, 5.3% (5.2% to 5.5%) for stage 3, 4.8% (4.6% to 5.0%) for stage 4 (Table 2). Advanced CKM syndrome occurred in 6.3% (5.9% to 6.7%) in the CNSCKD and 10.1% (9.9% to 10.4%) in the sixth CCDRFS (Table 3).

**Table 2.**
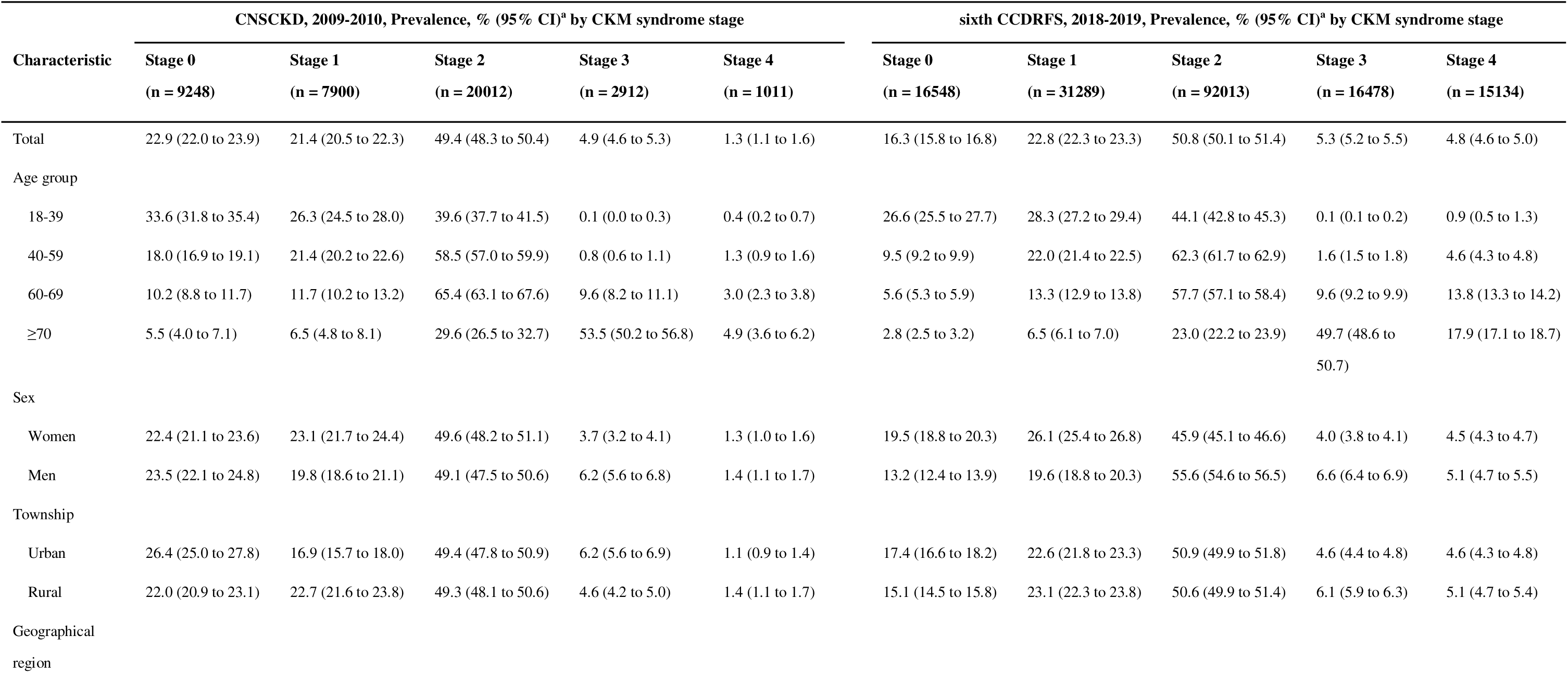
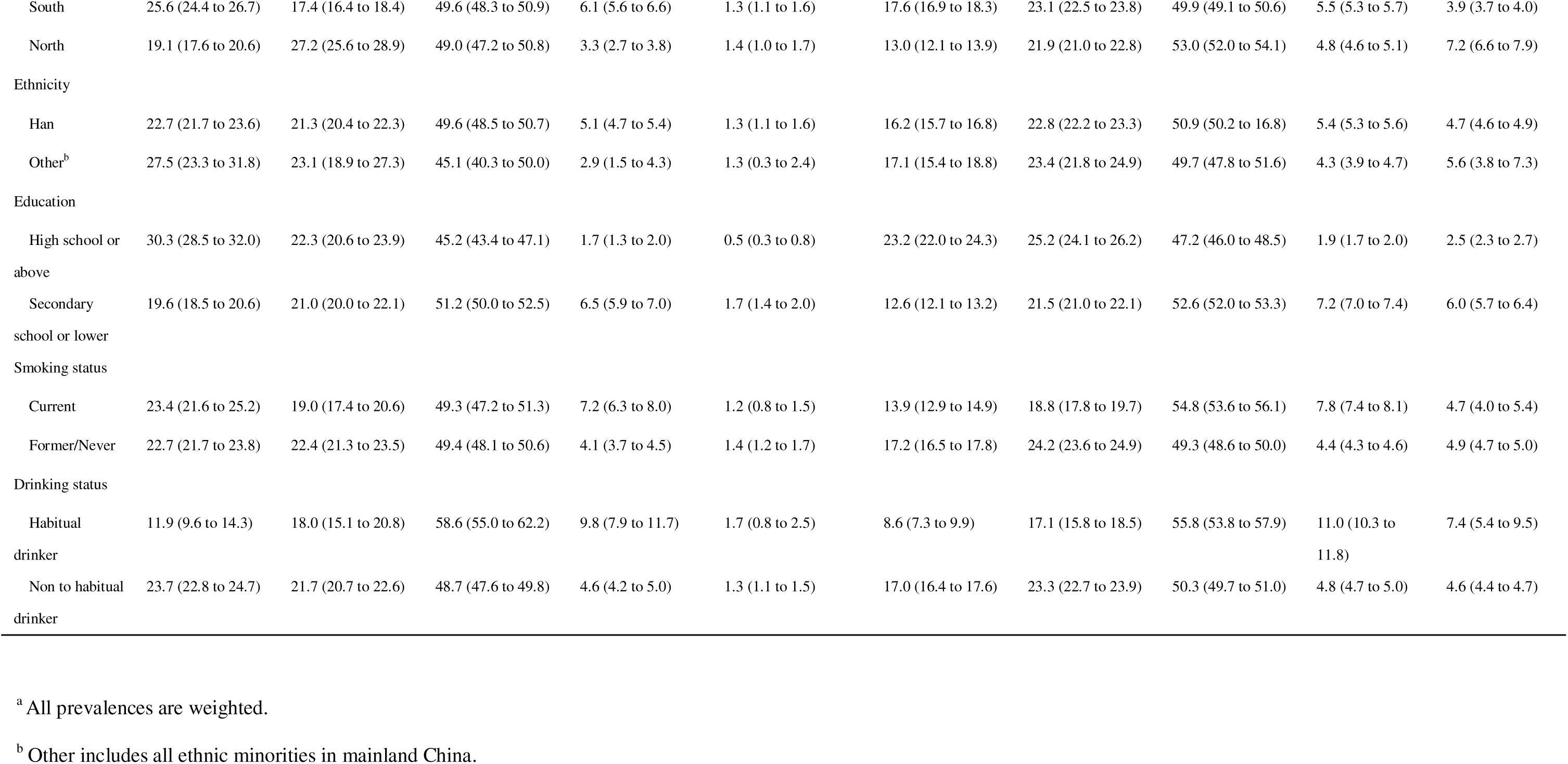
Weighted prevalence of cardiovascular-kidney-metabolic syndrome by stage in different strata among Chinese adults.

**Table 3.**
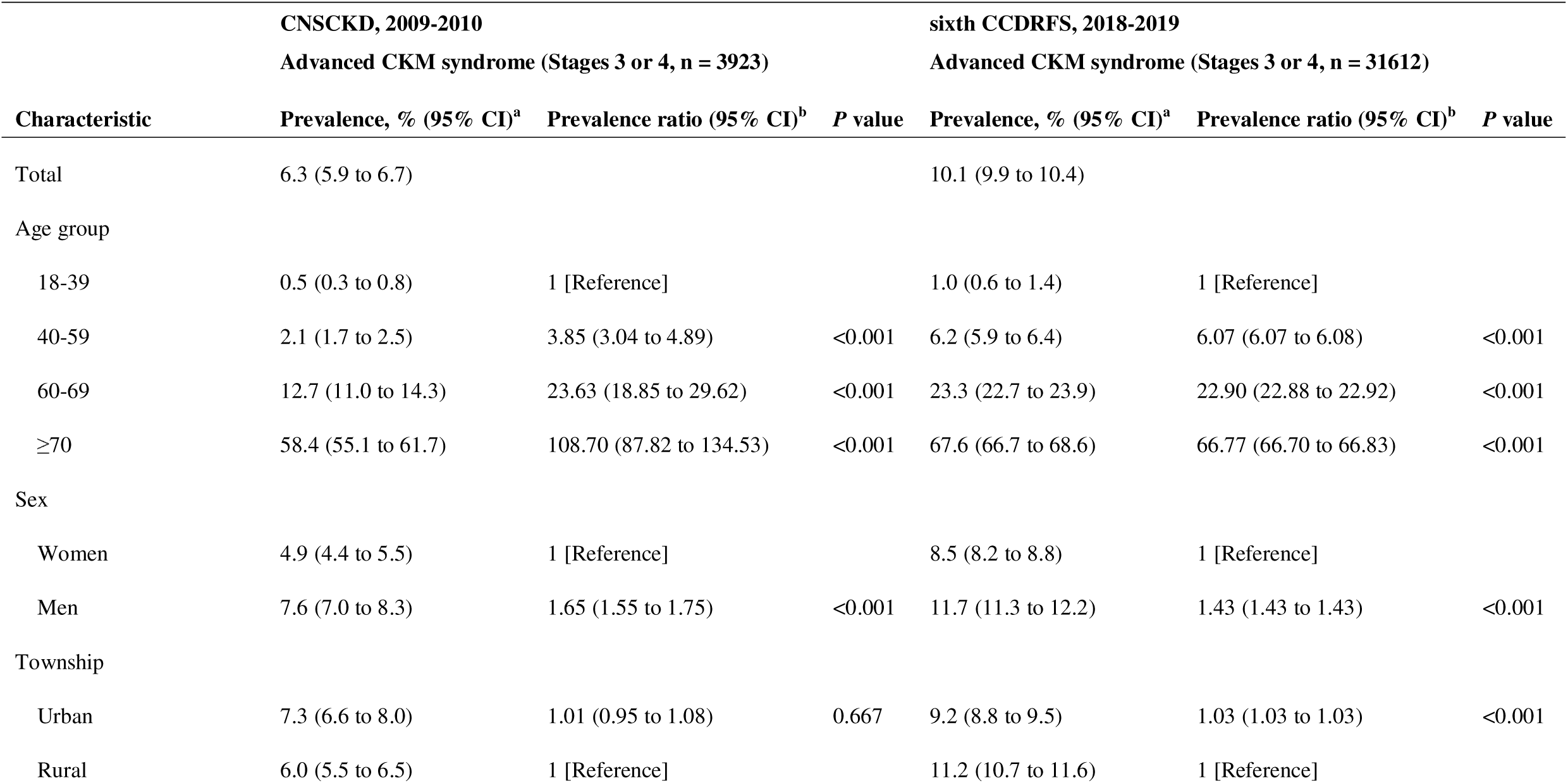
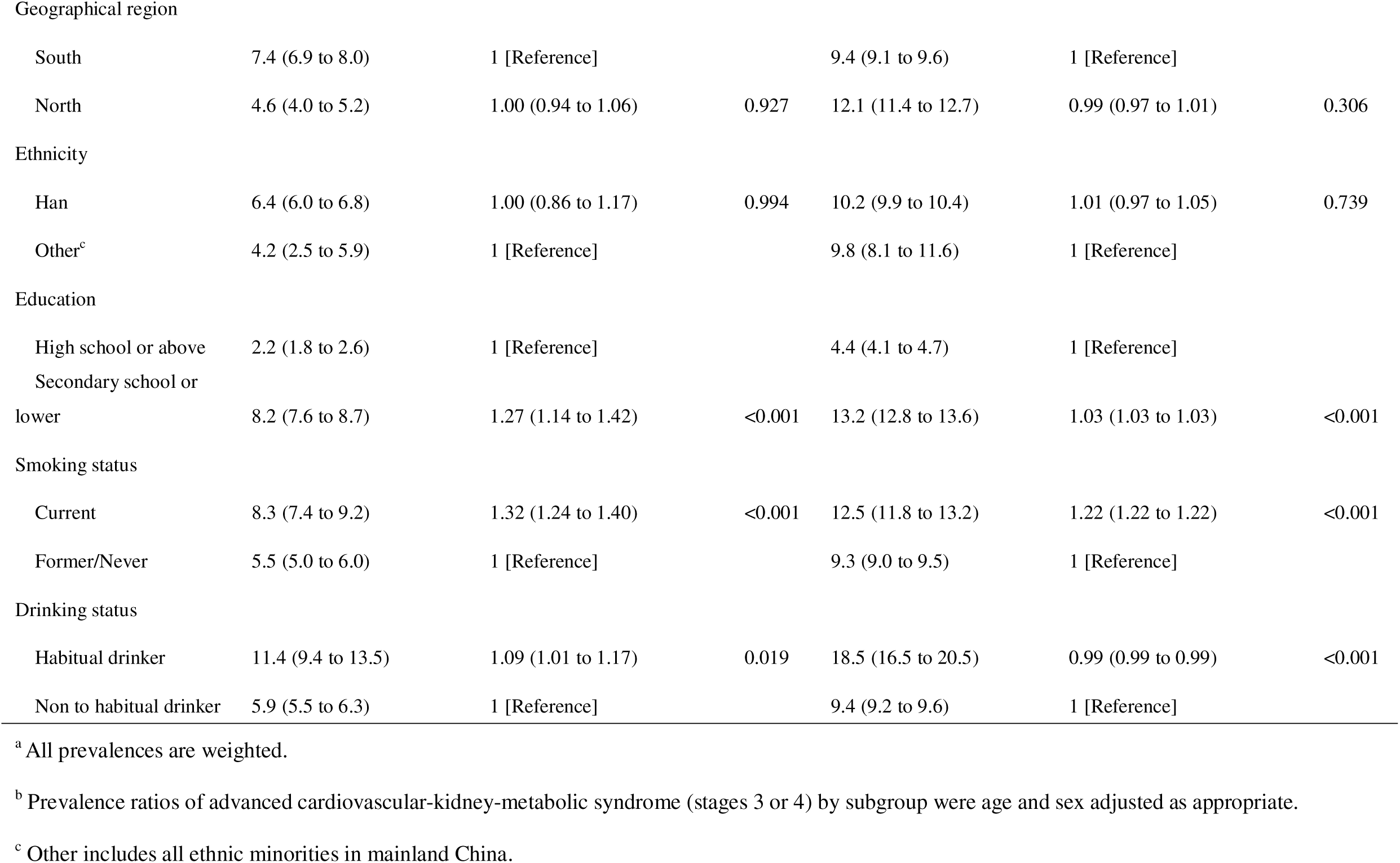
Weighted prevalence of advanced cardiovascular-kidney-metabolic syndrome (stages 3 or 4) in different strata among Chinese adults.

The prevalence of advanced CKM syndrome increased with age in both surveys and increased more sharply after age 60. Only 33.6% (31.8% to 35.4%) and 26.6% (25.5% to 27.7%) of adults aged 18-39 years had stage 0 in the CNSCKD and the sixth CCDRFS, respectively. Compared with women, men were more likely to have advanced CKM syndrome (7.6% vs 4.9%, adjusted prevalence ratio [aPR], 1.65 [95% CI, 1.55 to 1.75], *P* < 0.01 in the CNSCKD; 11.7% vs 8.5%, aPR, 1.43 [1.43 to 1.43], *P* < 0.001 in the sixth CCDRFS). When stratified by age and sex, the prevalence of stage 0 among males appears to have decreased by nearly half over the past decade, while in females, it has only shown a slight decline (supplementary table 2).

The prevalence of advanced CKM syndrome was higher among rural residents (the sixth CCDRFS only), people with a low education level, current smoker and habitual drinker. A higher prevalence of advanced CKM syndrome was also observed in the subgroups characterized by low fruit and vegetable intake, low red meat intake, and lacking physical activity in the sixth CCDRFS (supplementary table 3-4). No significant differences in the prevalence of advanced CKM syndrome stages were found between geographical regions or ethnicities.

### Potential change in distribution of risk factors in CKM syndrome stage 2

In the CKM stage 2 populations of both the two surveys, the highest weighted proportion was observed in individuals with only one risk factor, at 52.95% (51.49% to 54.41%) and 39.79% (38.84% to 40.74%), respectively (Figure 1).

**Figure 1.**
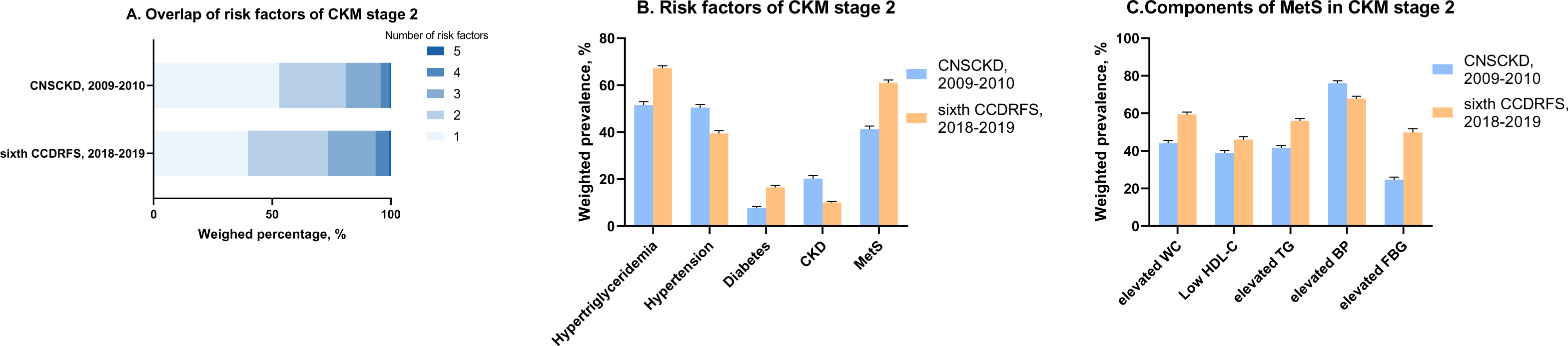
Weighted distribution of risk factors of cardiovascular-kidney-metabolic (CKM) syndrome stage 2 among Chinese adults. (A) Weighted distribution of numbers of risk factors of CKM syndrome stage 2; (B) Weighted distribution of risk factors of CKM syndrome stage 2; (C) Weighted distribution of components of metabolic syndrome in CKM stage 2. Abbreviations: CKD, chronic kidney disease; MetS, metabolic syndrome; WC, waist circumstance; HDL-C, high-density lipoprotein cholesterol; TG, triglyceride; BP, blood pressure; FBG, fasting blood glucose; CNSCKD, China National Survey of Chronic Kidney Disease; CCDRFS, China Chronic Disease and Risk Factor Surveillance.

In the CNSCKD, hypertriglyceridemia accounted for the highest weighted proportion (51.5%), followed by hypertension (50.4%) and metabolic syndrome (MetS, 41.2%). In the sixth CCDRFS, hypertriglyceridemia also had the highest weighted proportion (67.2%), followed by MetS (61.0%) and hypertension (39.4%).

The weighted proportions of MetS components also changed, with a decrease in the proportion of elevated blood pressure (76.0% in the CNSCKD; 67.8% in the sixth CCDRFS) and an increase in the proportions of abnormal WC (44.0% in the CNSCKD; 59.4% in the sixth CCDRFS) and elevated triglycerides (41.5% in the CNSCKD; 56.0% in the sixth CCDRFS).

### Factors associated with advanced CKM syndrome risk

Table 4 presents multivariable logistic regression results, where older age, male sex, urban residency, low education level, current smoking, and habitual alcohol consumption were significantly associated with an increased risk of advanced CKM syndrome. Residence in northern regions of China, Han ethnicity, low levels of fruit and vegetable intake, red meat intake, and physical activity were found to be associated with advanced CKM syndrome only in the sixth CCDRFS (supplementary table 5).

**Table 4.** Logistic regression analyses on advanced cardiovascular-kidney-metabolic syndrome (stages 3 or 4).

| Characteristic | CNSCKD, 2009-2010<br>Advanced CKM syndrome (Stages 3 or 4, n = 3923) |  | sixth CDRFS, 2018-2019<br>Advanced CKM syndrome (Stages 3 or 4, n = 31612) |  |
| --- | --- | --- | --- | --- |
|  | OR (95% CI)<sup>a</sup> | *P* value | OR (95% CI)<sup>a</sup> | *P* value |
| Age group |  |  |  |  |
| 18-39 | 1 [Reference] |  | 1 [Reference] |  |
| 40-59 | 3.50 (2.75 to 4.46) | <0.001 | 5.98 (5.97 to 5.99) | <0.001 |
| 60-69 | 23.04 (18.13 to 29.29) | <0.001 | 28.70 (28.67 to 28.73) | <0.001 |
| ≥70 | 261.28 (206.23 to 331.03) | <0.001 | 223.43 (223.18 to 223.68) | <0.001 |
| Sex |  |  |  |  |
| Women | 1 [Reference] |  | 1 [Reference] |  |
| Men | 2.08 (1.83 to 2.37) | <0.001 | 1.58 (1.58 to 1.59) | <0.001 |
| Township |  |  |  |  |
| Urban | 1.32 (1.14 to 1.51) | <0.001 | 1.11 (1.11 to 1.11) | <0.001 |
| Rural | 1 [Reference] |  | 1 [Reference] |  |
| Geographical region |  |  |  |  |
| South | 1 [Reference] |  | 1 [Reference] |  |
| North | 1.06 (0.94 to 1.20) | 0.363 | 1.70 (1.70 to 1.70) | <0.001 |
| Ethnicity |  |  |  |  |
| Han | 0.83 (0.62 to 1.11) | 0.208 | 0.92 (0.92 to 0.92) | <0.001 |
| Other<sup>b</sup> | 1 [Reference] |  | 1 [Reference] |  |
| Education |  |  |  |  |
| High school or above | 1 [Reference] |  | 1 [Reference] |  |
| Secondary school or lower | 1.74 (1.47 to 2.06) | <0.001 | 1.25 (1.25 to 1.25) | <0.001 |
| Smoking status |  |  |  |  |
| Current | 1.56 (1.36 to 1.78) | <0.001 | 1.52 (1.52 to 1.52) | <0.001 |
| Former/Never | 1 [Reference] |  | 1 [Reference] |  |
| Drinking status |  |  |  |  |
| Habitual drinker | 0.96 (0.81 to 1.15) | 0.671 | 1.07 (1.07 to 1.07) | <0.001 |
| Non to habitual drinker | 1 [Reference] |  | 1 [Reference] |  |

<sup>a</sup> The ORs were estimated from a single logistic model that included all the variables in the Table as covariates.
<sup>b</sup> Other includes all ethnic minorities in mainland China.

### Awareness and control of comorbidities

Prevalence, awareness, and control of comorbidities, including hypertension, diabetes, dyslipidemia, hyperuricemia and CKD in the individuals with CKM syndrome in the sixth CCDRFS are summarized in supplementary table 6. Among those with CKM stage 2, the weighted prevalence of dyslipidemia was the highest at 55.9%, followed by hypertension (39.4%), hyperuricemia (18.7%), diabetes (16.5%), and CKD (10.0%). The awareness rate of dyslipidemia and CKD was considerably lower than that of hypertension and diabetes.

## Discussion

In this large national population-based study, we found that 77.1% of adults aged 18 years or older in mainland China had CKM syndrome in 2010 and 83.7% had CKM syndrome in 2019, amounting to an estimated 875 million affected adults based on the sixth national census. Additionally, 6.3% of adults met the criteria for advanced CKM syndrome in 2010 and 10.1% in 2019.

A recent study involving 97 777 Chinese participants revealed almost 96% had CKM syndrome.(21) The discrepancy in prevalence may be attributed to the characteristics of the study population, which primarily involved middle-aged and elderly men from a northern Chinese community. Despite the rapid and sustained growth in the burden of CKM conditions in low- and middle-income countries, existing studies on the prevalence of CKM syndrome predominantly originated from high-income countries. Our findings indicate an upward trend in CKM syndrome prevalence over the past decade across all age groups, which aligns with previous studies on the prevalence of CKM components in China during the same period.(12) The estimated CKM syndrome prevalence in our study (83.7%) was slightly lower than that in the US (89.4%), mainly due to differences in the prevalence of advanced CKM syndrome (10.1% vs. 14.6%).(22) Notably, the prevalence of advanced CKM syndrome in China has increased by 60.3% over the past decade. Given the progressive nature and pathophysiological continuum of CKM syndrome, the prevalence in China is likely to continue rising without effective interventions, potentially closing the gap with the US over time.

Since the 1990s, China’s rapid urbanization triggered major changes in lifestyle and diet, driving a sharp rise in metabolic risk factors. These changes, initially concentrated in urban areas, have quickly spread to rural regions.(12) Despite this rise occurring over less than 40 years, the prevalence of CKM syndrome in China already approaches levels seen in the US.(22) This highlights an urgent need for effective early prevention strategies to address this growing health burden. China’s Basic Public Health Service program, launched in 2009, has contributed to mitigating non-communicable diseases, particularly through improvements in hypertension and diabetes management.(23) Building on this progress, kidney disease management should now be incorporated alongside existing efforts, with an integrated approach that targets all aspects of CKM syndrome to enhance early detection, intervention, and long-term health outcomes. Additionally, the prevalence of advanced CKM increased by 86.7% in rural areas between surveys, significantly exceeding the 26.0% increase in urban areas. This rural-urban disparity reflects gaps in healthcare resources and health literacy in rural regions, emphasizing the need for targeted public health strategies to reduce inequities and strengthen CKM prevention and care in underserved populations.

The notable increase in the prevalence of elevated WC between two surveys, particularly within CKM stage 2, highlights a concerning trend for future cardiovascular and metabolic health in China. Elevated WC is a key component of metabolic syndrome and is closely linked to abnormal glucose metabolism, insulin resistance, and systemic inflammation, all of which increase the risk of CKM progression.(24) This trend underscores the growing burden of metabolic risk factors in China and the need for targeted interventions to prevent further escalation. Monitoring and managing WC could play a pivotal role in reducing CKM-related health risks.

In addition to traditional CKM risk factors, our study suggests that social determinants of health (SDoH) and personal behaviors play a significant role in preventing advanced CKM syndrome. A study based on US county-level data found a significant negative association between CKM-related mortality and high school completion rates, highlighting the importance of improving SDoH at the population level.(25) Numerous studies have demonstrated that lifestyles are modifiable risk factors for adverse health outcomes, including diabetes and CVD.(26–28) Recently, a study in China involving 100 000 participants also emphasized the significance of healthy lifestyles, particularly for young individuals with high genetic risk.(29) Our findings support the need for public health programs and interventions aimed at improving health behaviors to reduce CKM syndrome risk.

To the best of our knowledge, this study is the first to examine CKM syndrome prevalence among Chinese adults using two nationally representative sample. Nevertheless, we also acknowledge some limitations. First, CVD history was self-reported, and certain data, such as cardiac biomarkers and cardiovascular imaging, recommended for defining advanced CKM syndrome stages, were unavailable, which may lead to an underestimation of advanced stages in both surveys. Second, because the definition of CVD history differed between the two surveys, caution should be exercised when comparing the prevalence of stage 4 across surveys. Third, differences in methods used to define CKD and diabetes between the two surveys could affect prevalence estimates. Nevertheless, the main results remained largely consistent across surveys.

In conclusion, the prevalence of CKM syndrome in China is high, and has increased significantly over the past decade, emphasizing the urgent need to develop comprehensive and equitable prevention and management strategies.

## Data Availability

Individual participant data will not be made available publicly. For further detailed data access policy and procedure, please contact with the corresponding authors.

## Contributors

DH, XZ, LW, LZ and ED developed the study concept and drafted analyses plan. DH, XZ, YC, MY, JW, BG, MZ, MZ, LW and LZ collected data. DH and XZ conducted analysis and prepared results. DH, XZ, LW, LZ and ED wrote the first draft of the paper. CL, JH, BG, MZ and MZ provided critical review. All authors provided input into interpretation of results and contents of the paper. LW and LZ had full access to all of the data in the study and verified the data. LW, LZ and ED are responsible for the integrity of the data, accuracy of the data analysis, and decision to submit the manuscript, and act as guarantors. The corresponding author attests that all listed authors meet authorship criteria and that no others meeting the criteria have been omitted.

## Declaration of interests

We declare no competing interests.

## Sources of Funding

This study was supported by grants from the National Natural Science Foundation of China (No. 72125009), National Key Research and Development Program of China (No. 2022YFF1203001), National Natural Science Foundation of China (No. 81771938, 81900665, 82090021), National Key R&D Program of the Ministry of Science and Technology of China (No.2021YFC2500201, No. 2019YFC2005000), Young Elite Scientists Sponsorship Program by CAST (No. 2022QNRC001), National High Level Hospital Clinical Research Funding (State Key Laboratory of Vascular Homeostasis and Remodeling, Peking University), Innovation Fund for Outstanding Doctoral Candidates of Peking University Health Science Center (No. BMU2024BSS001), and CAMS Innovation Fund for Medical Sciences (No. 2019-I2M-5-046).

## Acknowledgments

We thank all the survey teams and all the study participants in the CNSCKD and the sixth CCDRFS, for their contributions and dedication to this study.

## Notes

### Competing Interest Statement

The authors have declared no competing interest.

### Author Declarations

The study protocol was approved by the ethics review committee of the National Center for Chronic and Noncommunicable Diseases Control and Prevention, and Peking University First Hospital.

