## Supplementary Materials for "Rising Prevalence of Cardiovascular-Kidney-Metabolic (CKM) Syndrome in China, 2010-2019: National Cross-Sectional Surveys"

| **Supplementary Method 1.** | Data collection methods in the China National Survey of Chronic Kidney Disease (CNSCKD) and the sixth China Chronic Disease and Risk Factor Surveillance (CCDRFS). |
| --- | --- |
| **Supplementa****ry Table 1.** | Baseline characteristics of diet and physical activity among participants from the sixth China Chronic Disease and Risk Factor Surveillance (CCDRFS) according to cardiovascular-kidney-metabolic syndrome stages. |
| **Supplementary Table 2.** | Weighted prevalence of cardiovascular-kidney-metabolic syndrome by stage stratified by age and sex among Chinese adults. |
| **Supplementary Table 3.** | Weighted prevalence of cardiovascular-kidney-metabolic syndrome by stage in different strata of diet and physical activity among participants from the sixth China Chronic Disease and Risk Factor Surveillance (CCDRFS). |
| **Supplementary Table 4.** | Weighted prevalence of advanced cardiovascular-kidney-metabolic syndrome (stages 3 or 4) in different strata of diet and physical activity among participants from the sixth China Chronic Disease and Risk Factor Surveillance (CCDRFS). |
| **Supplementary Table 5.** | Logistic regression analyses on advanced cardiovascular-kidney-metabolic syndrome (stages 3 or 4) in the sixth China Chronic Disease and Risk Factor Surveillance (CCDRFS). |
| **Supplementary Table 6.** | Weighted prevalence, awareness, treatment, and control of comorbidities among those with different cardiovascular-kidney-metabolic syndrome stages, and total population in sixth CCDRFS, 2018-2019. |
| **Supplementary Figure 1.** | Participants selection diagram. |

**Supplemental Method 1. Data collection methods in the China National Survey of Chronic Kidney Disease (CNSCKD) and the sixth China Chronic Disease and Risk Factor Surveillance (CCDRFS).**

| **Covariates** | **CNSCKD, 2009-2010** | **sixth CCDRFS, 2018-2019** |
| --- | --- | --- |
| Blood pressure | Blood pressure was measured by sphygmomanometer, three times at 5 min intervals. The mean of the three readings was calculated, unless the difference between the readings was greater than 10 mmHg, in which case the mean of the two closest measurements was used. | Blood pressure was measured on the nondominant arm three times consecutively with a 1-minute interval between measurements while the participant was seated, after 5 minutes of rest, using an automated device. The average of the last two readings was used for analysis. |
| Creatinine | Serum and urinary creatinine were measured with Jaffe’s kinetic method. | Serum and urinal creatinine were measured by an enzymatic assay that was traceable to isotope dilution mass spectrometry. |
| eGFR | Estimated glomerular filtration rate (eGFR) was calculated with 2021 race-free Chronic Kidney Disease Epidemiology Collaboration (CKD-EPI) creatinine equation. | eGFR was calculated with 2021 race-free CKD-EPI creatinine equation. |
| uACR | Urinary albumin was measured with immunoturbidimetric tests. The urinary albumin to creatinine ratio (uACR; mg/g) was calculated, with albuminuria defined as a uACR of 30 mg/g or higher. | Urinary albumin was measured with immunoturbidimetric tests. The urinary albumin to creatinine ratio (uACR; mg/g) was calculated, with albuminuria defined as a uACR of 30 mg/g or higher. |
| Fasting blood glucose | Fasting blood glucose was measured enzymatically with a glucose oxidase method. | Fasting blood glucose was were measured using commercial kits on automatic analyzers. |
| HbA1c | - | HbA1c was directly measured from venous blood samples using quantitative high-performance liquid chromatography and the boronated affinity method on a hemoglobin analyzer (D-10 [Bio-Rad]). |
| Serum lipids | Serum lipids were measured with commercially available reagents in all laboratories. | Serum lipids were measured using commercial kits on automatic analyzers. |
| Waist circumference | Elevated waist circumference: ≥80 and ≥90 cm for women and men with Asian race, respectively. | Elevated waist circumference: ≥80 and ≥90 cm for women and men with Asian race, respectively. |
| BMI | BMI was calculated as weight in kilograms divided by height in meters squared. Elevated BMI: ≥23 kg/m^2^ for individuals with Asian race. | BMI was calculated as weight in kilograms divided by height in meters squared. Elevated BMI: ≥23 kg/m^2^ for individuals with Asian race. |
| Hypertension | Hypertension was defined as a systolic blood pressure (SBP) of 140 mmHg or more, diastolic blood pressure (DBP) of 90 mmHg, any use of antihypertensive medication, or any self-reported history of hypertension. | Hypertension was defined as a SBP of 140 mmHg or more, DBP of 90 mmHg, any use of antihypertensive medication, or any self-reported history of hypertension. |
| Prediabetes | Prediabetes was defined as a fasting blood glucose of 100 mg/dL to <126 mg/dL. | Prediabetes was defined as a HbA1c of 5.7% to <6.5% or a fasting blood glucose of 100 mg/dL to <126 mg/dL. |
| Diabetes | Diabetes was defined as a fasting plasma glucose of 126 mg/dL or more, any use of hypoglycemic agent, or any self-reported history of diabetes. | Diabetes was defined as a fasting plasma glucose of 126 mg/dL or higher, a 2-hour plasma glucose of 200 mg/dL or higher in oral glucose tolerance test, an HbA1c of 6.5% or higher, any use of hypoglycemic agent, or any self-reported history of diabetes. |
| Chronic kidney disease | Chronic kidney disease was defined as eGFR < 60 mL/min/1.73m^2^ or albuminuria. | Chronic kidney disease was defined as eGFR < 60 mL/min/1.73m^2^ or albuminuria. |
| Metabolic syndrome | MetS, ≥3 of the following: elevated WC, low high density lipoprotein cholesterol (HDL-C) level [<40 mg/dL or <50 mg/dL for men or women, respectively], TG ≥150 mg/dL, elevated blood pressure [systolic blood pressure (SBP) ≥130, diastolic blood pressure (DBP) ≥80 mmHg, and/or use of blood pressure-lowering medications], or fasting blood glucose ≥100 mg/dL | MetS, ≥3 of the following: elevated WC, low high density lipoprotein cholesterol (HDL-C) level [<40 mg/dL or <50 mg/dL for men or women, respectively], TG ≥150 mg/dL, elevated blood pressure [systolic blood pressure (SBP) ≥130, diastolic blood pressure (DBP) ≥80 mmHg, and/or use of blood pressure-lowering medications], or fasting blood glucose ≥100 mg/dL |
| CVD | myocardial infarction and stroke | myocardial infarction, angina, atrial fibrillation, and stroke |

**Supplementary Table 1. Baseline characteristics of diet and physical activity among participants from the sixth China Chronic Disease and Risk Factor Surveillance (CCDRFS) according to cardiovascular-kidney-metabolic syndrome stages.**

|  | sixth CCDRFS, 2018-2019, No. (%)^a^ | | | | | |
| --- | --- | --- | --- | --- | --- | --- |
| Characteristic | Total (n=171 462) | Stage 0 (n = 16 548) | Stage 1 (n = 31 289) | Stage 2 (n = 92 013) | Stage 3 (n = 16 478) | Stage 4 (n = 15 134) |
| Fruit/vegetable intake |  |  |  |  |  |  |
| <400 g/d | 78412 (44.5) | 7566 (44.4) | 13989 (42.3) | 41000 (43.8) | 8428 (53.9) | 7429 (50.1) |
| ≥400 g/d | 88651 (55.5) | 8594 (55.6) | 16534 (57.1) | 48559 (56.2) | 7626 (46.1) | 7338 (49.9) |
| Red meat intake |  |  |  |  |  |  |
| ≥100 g/d | 60385 (42.2) | 6689 (46.2) | 12202 (43.7) | 32865 (43.1) | 5104 (31.1) | 3525 (23.8) |
| <100 g/d | 106678 (57.8) | 9471 (53.8) | 18321 (56.3) | 56694 (56.9) | 10950 (68.9) | 11242 (76.2) |
| Physical inactivity |  |  |  |  |  |  |
| <150 min/week | 33349 (22.0) | 3094 (22.4) | 5406 (19.6) | 17133 (22.2) | 4264 (26.7) | 3452 (24.7) |
| ≥150 min/week | 137509 (78.0) | 13419 (77.6) | 25794 (80.4) | 74547 (77.8) | 12139 (73.3) | 11610 (75.3) |

^a^ All percentages are weighted.

**Supplementary Table 2. Weighted prevalence of cardiovascular-kidney-metabolic syndrome by stage stratified by age and sex among Chinese adults.**

|  | CNSCKD, 2009-2010 Prevalence, % (95% CI)^a^ by cardiovascular-kidney-metabolic syndrome stage | | | | | sixth CCDRFS, 2018-2019 Prevalence, % (95% CI) ^a^ by cardiovascular-kidney-metabolic syndrome stage | | | | |
| --- | --- | --- | --- | --- | --- | --- | --- | --- | --- | --- |
| Characteristic | Stage 0 | Stage 1 | Stage 2 | Stage 3 | Stage 4 | Stage 0 | Stage 1 | Stage 2 | Stage 3 | Stage 4 |
| 18-39 |  |  |  |  |  |  |  |  |  |  |
| Men | 33.8 (31.2 to 36.3) | 22.6 (20.4 to 24.9) | 43.0 (40.4 to 45.7) | 0.1 (0.0 to 0.2) | 0.5 (0.1 to 0.9) | 19.9 (18.3 to 21.4) | 23.1 (21.6 to 24.7) | 55.7 (53.8 to 57.6) | 0.1 (0.1 to 0.2) | 1.2 (0.4 to 1.9) |
| Women | 33.5 (30.9 to 36.0) | 30.2 (27.6 to 32.8) | 35.9 (33.2 to 38.5) | 0.2 (0.0 to 0.5) | 0.3 (0.0 to 0.6) | 33.7 (32.1 to 35.2) | 33.7 (32.2 to 35.2) | 31.9 (30.5 to 33.4) | 0.1 (0.02 to 0.2) | 0.6 (0.4 to 0.9) |
| 40-59 |  |  |  |  |  |  |  |  |  |  |
| Men | 17.7 (16.0 to 19.4) | 20.7 (18.9 to 22.5) | 58.8 (56.7 to 61.0) | 1.2 (0.7 to 1.6) | 1.6 (1.0 to 2.1) | 8.6 (8.1 to 9.2) | 18.8 (18.1 to 19.6) | 65.0 (64.1 to 65.9) | 2.6 (2.3 to 2.8) | 5.0 (4.6 to 5.3) |
| Women | 18.3 (16.8 to 19.8) | 22.1 (20.5 to 23.8) | 58.1 (56.2 to 60.1) | 0.4 (0.2 to 0.7) | 1.0 (0.6 to 1.4) | 10.4 (9.9 to 10.9) | 25.2 (24.4 to 25.9) | 59.6 (58.8 to 60.4) | 0.7 (0.6 to 0.8) | 4.1 (3.8 to 4.4) |
| 60-69 |  |  |  |  |  |  |  |  |  |  |
| Men | 13.4 (11.0 to 15.8) | 13.5 (11.1 to 15.9) | 55.7 (52.2 to 59.2) | 14.7 (12.1 to 17.3) | 2.7 (1.7 to 3.7) | 7.0 (6.5 to 7.5) | 15.2 (14.4 to 15.9) | 50.0 (49.0 to 51.0) | 13.8 (13.2 to 14.5) | 14.0 (13.3 to 14.7) |
| Women | 7.2 (5.7 to 8.7) | 10.0 (8.3 to 11.7) | 74.8 (72.2 to 77.4) | 4.7 (3.3 to 6.0) | 3.4 (2.2 to 4.5) | 4.2 (3.9 to 4.6) | 11.4 (10.9 to 12.0) | 65.6 (64.7 to 66.5) | 5.2 (4.8 to 5.6) | 13.5 (12.9 to 14.2) |
| ≥70 |  |  |  |  |  |  |  |  |  |  |
| Men | 6.1 (3.8 to 8.4) | 5.5 (3.5 to 7.4) | 16.3 (12.8 to 19.8) | 67.7 (63.4 to 72.1) | 4.4 (2.7 to 6.1) | 2.8 (2.4 to 3.2) | 6.4 (5.7 to 7.1) | 11.5 (10.6 to 12.3) | 60.6 (59.1 to 62.0) | 18.7 (17.5 to 19.9) |
| Women | 5.1 (3.0 to 7.1) | 7.4 (4.7 to 10.0) | 41.5 (36.9 to 46.2) | 40.7 (36.1 to 45.3) | 5.4 (3.3 to 7.4) | 2.9 (2.4 to 3.4) | 6.7 (6.0 to 7.4) | 33.4 (32.0 to 34.7) | 39.9 (38.4 to 41.4) | 17.2 (16.1 to 18.3) |

^a^ All prevalences are weighted.

**Supplementary Table 3. Weighted prevalence of cardiovascular-kidney-metabolic syndrome by stage in different strata of diet and physical activity among participants from the sixth China Chronic Disease and Risk Factor Surveillance (CCDRFS).**

|  | sixth CCDRFS, 2018 to 2019, Prevalence, % (95% CI)^a^ by CKM syndrome stage | | | | |
| --- | --- | --- | --- | --- | --- |
| Characteristic | Stage 0 (n = 16 548) | Stage 1 (n = 31 289) | Stage 2 (n = 92 013) | Stage 3 (n = 16 478) | Stage 4 (n = 15 134) |
| Total | 16.3 (15.8 to 16.8) | 22.8 (22.3 to 23.3) | 50.8 (50.1 to 51.4) | 5.3 (5.2 to 5.5) | 4.8 (4.6 to 5.0) |
| Fruit/vegetable intake |  |  |  |  |  |
| <400 g/d | 16.3 (15.4 to 17.1) | 22.0 (21.2 to 22.9) | 49.8 (48.8 to 50.8) | 6.4 (6.2 to 6.7) | 5.4 (5.0 to 5.8) |
| ≥400 g/d | 16.4 (15.7 to 17.1) | 23.6 (22.9 to 24.3) | 51.3 (50.5 to 52.1) | 4.4 (4.3 to 4.6) | 4.3 (4.1 to 4.6) |
| Red meat intake |  |  |  |  |  |
| ≥100 g/d | 17.9 (17.0 to 18.8) | 23.7 (22.8 to 24.6) | 51.7 (50.7 to 52.8) | 3.9 (3.7 to 4.1) | 2.7 (2.5 to 2.9) |
| <100 g/d | 15.2 (14.5 to 15.9) | 22.3 (21.6 to 23.0) | 49.8 (49.0 to 50.6) | 6.3 (6.1 to 6.5) | 6.4 (6.0 to 6.7) |
| Physical inactivity |  |  |  |  |  |
| <150 min/week | 16.6 (15.4 to 17.9) | 20.4 (19.3 to 21.5) | 51.2 (49.7 to 52.6) | 6.4 (6.1 to 6.8) | 5.4 (4.6 to 6.2) |
| ≥150 min/week | 16.2 (15.6 to 16.8) | 23.5 (22.9 to 24.1) | 50.6 (49.9 to 51.3) | 5.0 (4.8 to 5.1) | 4.6 (4.5 to 4.8) |

^a^ All prevalences are weighted.

**Supplementary Table 4. Weighted prevalence of advanced cardiovascular-kidney-metabolic syndrome (stages 3 or 4) in different strata of diet and physical activity among participants from the sixth China Chronic Disease and Risk Factor Surveillance (CCDRFS).**

|  | Advanced CKM syndrome (Stages 3 or 4, n = 31 612) | | |
| --- | --- | --- | --- |
| Characteristic | Prevalence, % (95% CI)^a^ | Prevalence ratio (95% CI)^b^ | *P* value |
| Total | 10.1 (9.9 to 10.4) |  |  |
| Fruit/vegetable intake |  |  |  |
| <400 g/d | 11.9 (11.4 to 12.3) | 1.05 (1.05 to 1.05) |  |
| ≥400 g/d | 8.8 (8.5 to 9.0) | 1 [Reference] | <0.001 |
| Red meat intake |  |  |  |
| ≥100 g/d | 6.7 (6.4 to 6.9) | 0.88 (0.88 to 0.88) | <0.001 |
| <100 g/d | 12.7 (12.3 to 13.1) | 1 [Reference] |  |
| Physical inactivity |  |  |  |
| <150 min/week | 11.8 (11.0 to 12.6) | 1.10 (1.10 to 1.10) | <0.001 |
| ≥150 min/week | 9.6 (9.4 to 9.9) | 1 [Reference] |  |

^a^ All prevalences are weighted.

^b^ Prevalence ratios of advanced cardiovascular-kidney-metabolic syndrome (stages 3 or 4) by subgroup were age and sex adjusted as appropriate.

**Supplementary Table 5. Logistic regression analyses on advanced cardiovascular-kidney-metabolic syndrome (stages 3 or 4) in the sixth China Chronic Disease and Risk Factor Surveillance (CCDRFS).**

|  | Advanced CKM syndrome (Stages 3 or 4, n = 31612) | |
| --- | --- | --- |
| Characteristic | OR (95% CI)^a^ | *P* value |
| Age group |  |  |
| 18-39 | 1 [Reference] |  |
| 40-59 | 5.84 (5.83 to 5.84) | <0.001 |
| 60-69 | 27.40 (27.37 to 27.43) | <0.001 |
| ≥70 | 207.70 (207.47 to 207.95) | <0.001 |
| Sex |  |  |
| Women | 1 [Reference] |  |
| Men | 1.60 (1.60 to 1.60) | <0.001 |
| Township |  |  |
| Urban | 1.16 (1.16 to 1.16) | <0.001 |
| Rural | 1 [Reference] |  |
| Geographical region |  |  |
| South | 1 [Reference] |  |
| North | 1.60 (1.60 to 1.60) | <0.001 |
| Ethnicity |  |  |
| Han | 0.87 (0.87 to 0.87) | <0.001 |
| Other | 1 [Reference] |  |
| Education |  |  |
| High school or above | 1 [Reference] |  |
| Secondary school or lower | 1.20 (1.20 to 1.20) | <0.001 |
| Smoking status |  |  |
| Current | 1.57 (1.56 to 1.57) | <0.001 |
| Former/Never | 1 [Reference] |  |
| Drinking status |  |  |
| Habitual drinker | 1.11 (1.10 to 1.11) | <0.001 |
| Non to habitual drinker | 1 [Reference] |  |
| Fruit/vegetable intake |  |  |
| <400 g/d | 1 [Reference] |  |
| ≥400 g/d | 1.08 (1.08 to 1.08) | <0.001 |
| Red meat intake |  |  |
| ≥100 g/d | 0.71 (0.71 to 0.71) | <0.001 |
| <100 g/d | 1 [Reference] |  |
| Physical inactivity |  |  |
| <150 min/week | 1.21 (1.21 to 1.21) | <0.001 |
| ≥150 min/week | 1 [Reference] |  |

^a^ The ORs were estimated from a single logistic model that included all the variables in the Table as covariates.

**Supplementary Table 6. Weighted prevalence, awareness, treatment, and control of comorbidities among those with different cardiovascular-kidney-metabolic syndrome stages, and total population in sixth CCDRFS, 2018-2019.**

| **Comorbidity** | **Prevalence to % (95% CI)^a^** | **Awareness to % (95% CI)^a^** | **Treatment to % (95% CI)^a^** | **Control in treated patients to % (95% CI)^a^** |
| --- | --- | --- | --- | --- |
| Hypertension |  |  |  |  |
| Total population | 27.5 (26.6 to 28.4) | 40.6 (39.2 to 42.1) | 34.5 (33.1 to 35.8) | 32.2 (30.5 to 33.9) |
| CKM stage 2 | 39.4 (38.1 to 40.6) | 31.6 (30.1 to 33.1) | 25.4 (24.0 to 26.7) | 37.0 (35.1 to 38.9) |
| CKM stage 3 | 80.9 (79.8 to 82.0) | 58.1 (56.3 to 59.9) | 51.8 (49.9 to 53.8) | 22.3 (20.1 to 24.5) |
| CKM stage 4 | 66.7 (65.1 to 68.2) | 73.4 (71.5 to 75.4) | 67.9 (65.7 to 70.2) | 31.1 (28.5 to 33.7) |
| Diabetes |  |  |  |  |
| Total population | 12.4 (11.8 to 13.0) | 36.6 (34.6 to 38.5) | 32.8 (30.9 to 34.7) | 50.3 (47.6 to 53.0) |
| CKM stage 2 | 16.5 (15.5 to 17.4) | 31.7 (29.3 to 34.1) | 28.1 (25.7 to 30.4) | 50.2 (46.0 to 54.3) |
| CKM stage 3 | 49.6 (47.7 to 51.4) | 40.4 (38.3 to 42.5) | 36.2 (34.1 to 38.3) | 50.3 (47.1 to 53.6) |
| CKM stage 4 | 29.9 (28.4 to 31.3) | 57.9 (55.1 to 60.8) | 54.0 (51.2 to 56.7) | 50.6 (46.8 to 54.4) |
| Dyslipidemia |  |  |  |  |
| Total population | 38.4 (37.4 to 39.5) | 17.5 (16.4 to 18.6) | 10.2 (9.4 to 10.9) | 38.3 (35.8 to 40.8) |
| CKM stage 2 | 55.9 (54.5 to 57.2) | 14.9 (13.9 to 15.9) | 8.4 (7.7 to 9.0) | 31.2 (28.5 to 34.0) |
| CKM stage 3 | 47.2 (45.5 to 49.0) | 24.3 (22.4 to 26.3) | 13.4 (11.8 to 15.0) | 37.5 (32.1 to 42.9) |
| CKM stage 4 | 52.8 (50.7 to 55.0) | 47.1 (44.4 to 49.9) | 33.8 (31.2 to 36.5) | 45.3 (39.4 to 51.1) |
| Hyperuricemia^b^ |  |  |  |  |
| Total population | 14.2 (13.3 to 15.1) | - | - | - |
| CKM stage 2 | 18.7 (17.5 to 19.9) | - | - | - |
| CKM stage 3 | 19.3 (17.8 to 20.9) | - | - | - |
| CKM stage 4 | 12.8 (11.6 to 13.9) | - | - | - |
| Chronic kidney disease^c^ |  |  |  |  |
| Total population | 7.7 (7.3 to 8.1) | 9.9 (8.9 to 11.0) | - | - |
| CKM stage 2 | 10.0 (9.4 to 10.5) | 8.0 (6.9 to 9.1) | - | - |
| CKM stage 3 | 32.7 (31.2 to 34.2) | 13.0 (11.3 to 14.6) | - | - |
| CKM stage 4 | 18.3 (16.6 to 20.0) | 15.0 (12.3 to 17.8) | - | - |

^a^ All results are weighted.

^b^ Awareness, treatment, and control of hyperuricemia were not estimated because self-reported physician-diagnosed hyperuricemia was not ascertained.

^c^ Treatment and control of chronic kidney disease were not estimated because relevant information was lacking.

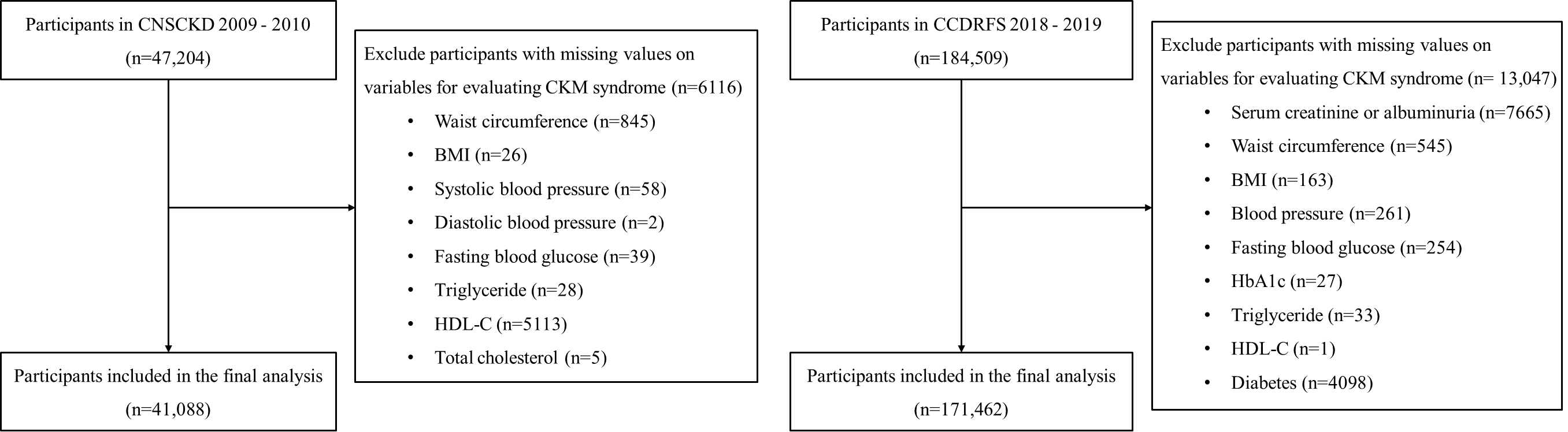

**Supplementary Figure 1. Participants selection diagram.**
